# Retrospective Analysis of the Utility of Genetic Testing in Predicting Drug Response in Chronic Pain

**DOI:** 10.1101/2021.04.29.21256336

**Authors:** Gaurav Gupta, Paquet-Proulx Cpl Emilie, Sasha Lalonde, Kira Burton, Besemann LCol Markus, Minerbi Amir

## Abstract

**Introduction:** Chronic pain is often unrecognized and/or undertreated, and as a result has significant impact on functional abilities, quality of life, societal participation and health care utilization. Medications remain a mainstay of treatment, but selection for any given patient remains a challenge when trying to predict efficacy and/or side effects. There is interest to see whether genetic analysis of how a given drug is processed for a patient can help with rational drug choices. This appears to have some early support in cardiac, psychiatric and acute pain studies. We studied whether genetic analysis of drug processing using the Pillcheck program could have helped in choosing the appropriate medications in a cohort of patients suffering from chronic pain. To our knowledge this type of study has not been completed in this environment and/or patient population.

**Methods:** We retrospectively studied a 31 patient cohort seen in the Canadian Forces Health Services Unit (Ottawa) Physiatry clinic. All patients suffered from a diagnosed chronic pain condition, completed the Pillcheck genetic drug processing analysis and filled-in questionnaires looking at efficacy and side effects of the drugs. We analyzed the correlation between the Pillcheck predictions and participants’ self-reported treatment efficacy and tolerability. The goal was to explore the clinical utility of Pillcheck results in guiding prescriptions for chronic pain patients.

**Results:** 31 patients returned completed questionnaires and had samples taken. Forty eight percent of the participants were actively treated with one of the study pain medications, and 84% had been taking at least one of these medications and discontinued. Pillcheck scores did not correlate with self-reported efficacy of any of the medications, nor did it correlate with self-reported side effects. Furthermore, active medications were more likely to receive a score indicating caution should be exerted, than were medications which had been discontinued.

**Discussion:** In a small cohort of pain patients with comorbid psychiatric disorders, genetic profiling using Pillcheck did not seem to correlate with reported benefit or side effect profile of commonly prescribed pain medications. Furthermore, discontinued medications were no more likely to be marked as warranting caution than did actively used medications.

**Conclusion:** Retrospectively using pharmacogenetics to guide medication selection in Canadian Forces members with chronic pain did not correlate with response or side effects. A larger prospective controlled study, measuring numerous clinical and non- clinical outcomes would be worthwhile in the future before widespread adoption for patients living with chronic pain.

## Introduction

Up to 20% of Canadians suffer from chronic pain with much higher estimated prevalence rates in older adults and those in long term care [1,2,3]. Chronic pain is often unrecognized and/or undertreated, and as a result has significant impact on functional abilities, quality of life, societal participation and health care utilization [4,5,6,7,8]. Even when compared with other diseases, quality of life in patients suffering from chronic pain is poorer [4], with associated productivity costs and health expenditures being up to $60 billion in Canada, and $635 billion in the United States per year [9,10].

While epidemiologic information regarding Canadian civilians and chronic pain exists [8], information regarding the Canadian Armed Forces (CAF) is limited. In Canadian Armed Forces veterans, 41% of the population experienced constant pain and 23% experienced intermittent chronic pain [11]. When comparing to the Canadian civilian population, the prevalence of activity reduction for veterans was higher (49% versus 21%), and a greater percentage required assistance with at least one activity of daily living (17% versus 5%) [12]. Disability odds were higher for chronic pain (10.9, p<.05), than mental health conditions (2.7 p<0.05), but not surprisingly there was a combined impact of physical and mental health issues [12]. In the Unites States, certain veteran populations have a chronic pain prevalence of about 47%, with moderate to severe pain in 28% [13]. Therefore, mental health and musculoskeletal/chronic pain issues are a leading cause of functional limitations, decreased quality of life, and economic loss in patients leaving military service [14].

Increasing knowledge of the mechanisms and factors related to the multidimensional nature of pain has translated into improved understanding of optimal care for the chronic pain patient. As a result, there has been an evolution in treatment approaches to pain including improved surgeries, interventional procedures, medications, psychology interventions, physical therapy, team-based approaches, and complementary approaches. Despite this, considerably more needs to be understood about which treatments are best for which patient and the efficacy and safety of various treatments over time.

Medications remain a mainstay of treatment, but selection for any given patient remains a challenge when trying to predict efficacy and/or side effects. The efficacy and side effects profile of a given medication is highly variable and dependent on multiple inherent and environmental factors, including genetics, interactions with the gut microbiome and drug-drug interactions to name just a few [15,16]. There is interest to see whether genetic analysis of how a given drug is processed for a patient can help with rational drug choices (i.e. most efficacious, with least side effects/interactions, in the fastest time). This appears to have some early support in cardiac, psychiatric and acute pain studies. In fact the US Federal Drug Administration have made dosing recommendations to included CPY2D6 and CYP2C19 polymorphisms for certain psychotropic medications [17]. There are numerous commercially available pharmacogenomic tests, however the data is limited regarding clinical utility based on existing studies and concerns regarding standardization between tests [18,19,20,21,22].

Metabolic processing of medication through the cytochrome enzymatic P450 system can vary depending, among other factors, on the allele’s present, and can lead to variable levels of clinical effect and/or side effects. In the case of opioids specifically this can mean a range as significant as decreased analgesia, through to life-threatening toxicity and death depending on the population present [23,24,25]. Smith et al showed a 24% of patients achieved a 30% reduction in pain when using the guided treatment decisions, for tramadol and codeine compared to patients in the usual care group for looking at immediate and poor metabolizers (8.3% of the cohort), with no difference/benefit noted for normal metabolizers [25].

Similarly, Greden *et al*. showed that patients taking medication for major depression disorder in congruence with their pharmacogenetic testing had a 30% higher rate of treatment response remission, but not symptom change, and a 50% improvement in remission compared to treatment as usual [26]. Alternately, Walden *et al*. recorded clinicians’ perspectives on prescribing using the pharmacogenetic testing. Of respondents discussing their patient’s outcomes 23% found utility, 41% reported no change, and no patients had a worsening of outcomes. Furthermore, patients with various metabolism profiles did not report any differences in side effects [17].

With respect to the healthcare resource utilization, one study found not using genetic information for prescribing had more medical visits, medical absence days, and disability claims [27]. Similarly, Elliot *et al*. in their single center randomized control trial showed with guided therapy that there was a 60 day relative risk decrease of 0.37 for re-hospitalization and 0.27 for emergency room visits [28]. Olson *et al*. showed using guided therapy decisions only 28% of patients (vs 53% in the control) experienced side effects, which was estimated to translate into a $1,100 of health care costs [29].

Understanding the diversity of metabolic potential while analyzing any given patient’s medication profile, is postulated to aid rational, precision drug choices. In the current study, we focused on whether genetic analysis of drug processing using the Pillcheck program could have helped in choosing the appropriate medications for a cohort of patients suffering from chronic pain. To our knowledge this type of study has not been completed in this environment and/or patient population.

## Methods

We recruited 31 participants who were Canadian Forces members between the ages of 18-60 years old, suffering from chronic pain, seen within the Canadian Forces Ottawa Physical Medicine and Rehabilitation clinic (CFO-P).

Participants enrolled in the study provided consent, and a buccal cell sample for CYP2D6 genotyping, with genotype results reported in their electronic health record (EHR). If permission was granted pharmacists provided recommendations to physicians by report and according to the CPIC guidelines for a wide range of medications, including medications for pain and mental health issues. In this study we looked specifically at our commonly prescribed medications including nortriptyline, amitriptyline, duloxetine, tramadol, tapentadol, buprenorphine patch and Cesamet. Classification of inferred phenotypes (i.e. ultrarapid, normal, intermediate and poor metabolizer) was consistent with the recently published guidance for allele function status) [21]. There were 3 possible scores for each tested medication, ranging 1-3, which included; 1) standard precaution; 2) caution - frequent monitoring; 3) caution - consider alternatives.

Patients were asked to retrospectively note and evaluate pain medications they had previously taken. To provide context they were given a list of previous medications used, date last prescribed and a picture of the pills. Questionnaires included perceived efficacy, severity of side effects, types of side effects, and general comments.

### Data analysis and statistical considerations

The distribution of Pillcheck predictive score was separately evaluated for medications actively taken by participants and for medications that had been discontinued. The distribution of prediction scores was compared using ANOVA. Pillcheck prediction scores were separately compared against participants reported efficacy and side effects profile using Spearman’s correlation coefficient, and the respective p values were calculated. As no single comparison reached statistical significance correction for multiple comparisons was not implemented.

## Results

31 patients returned completed questionnaires and had samples taken. Forty eight percent of the participants were actively treated with one of the study pain medications, and 84% had been taking at least one of these medications and discontinued use. The distribution of active and discontinued pharmaceutical treatment is presented in Table 1.

**Table 1:** Number of participants actively prescribed, or having discontinued treatment with each of the six study medications.

| medication | active treatment | discontinued treatment |
| --- | --- | --- |
| nortriptyline | 3 | 12 |
| amitriptyline | 0 | 11 |
| duloxetine | 3 | 15 |
| tramadol | 6 | 17 |
| nabilone | 5 | 2 |
| buprenorphine patch | 2 | 0 |

Pillcheck prediction scores, ranging 1-3 (see methods), were compared between medications which were actively taken (19) and medications which had been discontinued (57). Interestingly, active medications were more likely to receive a score of 2 or 3 (indicating caution should be exerted when these are prescribed), than were discontinued medications (Figure 1). This trend failed to reach statistical significance.

**Figure 1:**
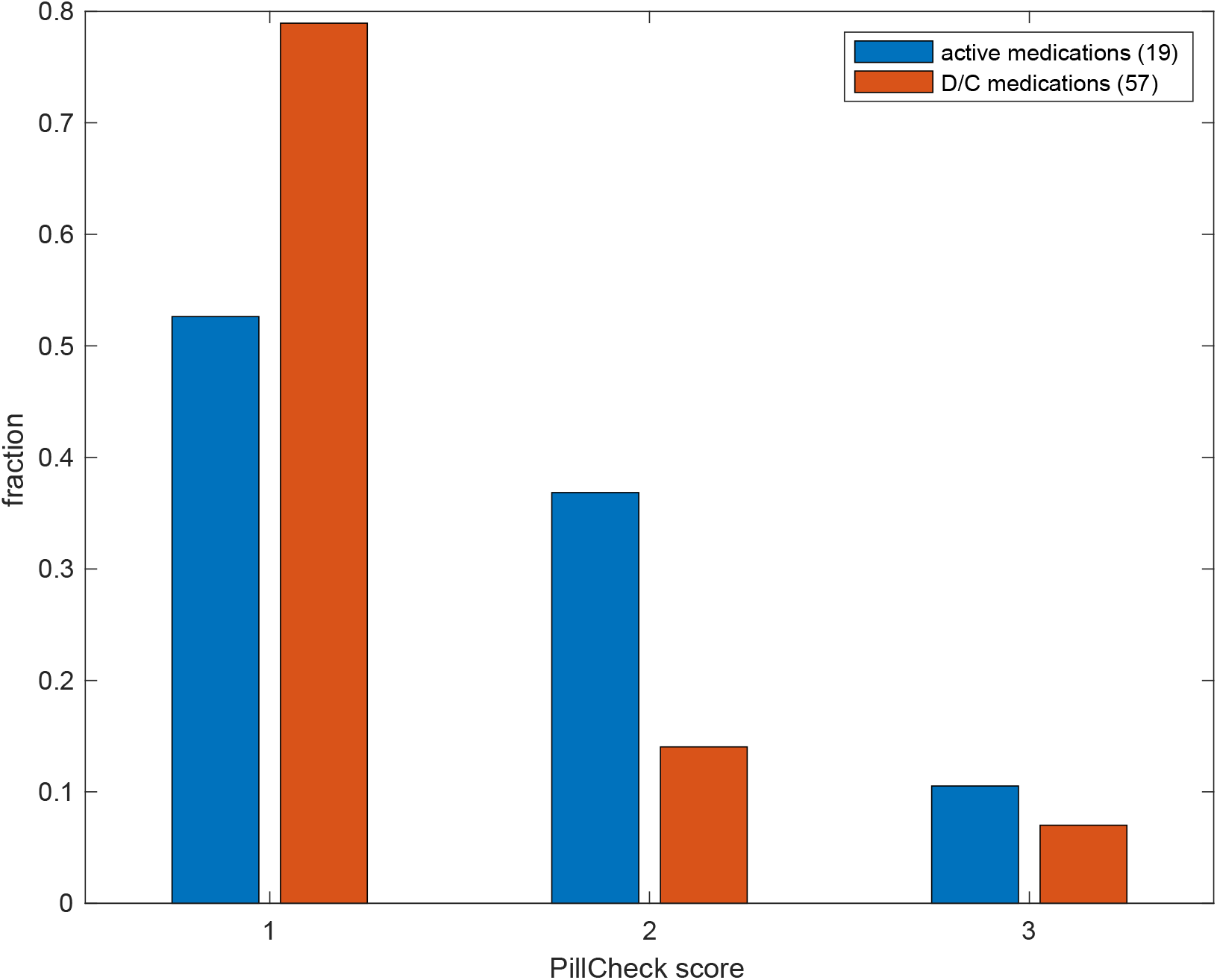
The distribution of Pillcheck prediction scores for actively prescribed (blue) and discontinued medications (orange); ANOVA p=0.07.

Consequently, Pillcheck scores were compared to self-reported efficacy and side effects profile, using Spearman’s correlations (Table 2). Of the six medications explored, 2 were excluded from this analysis as their Pillcheck score was identical (1), making the variance 0 and precluding a meaningful correlation analysis. Of the remaining 4 medications, none reached statistical significance, however a trend towards a negative correlation was observed for Cesamet, meaning that the higher the Pillcheck caution index was, the less side effects were reported by participants.

**Table 2:** Spearman’s correlation of Pillcheck prediction score and patient self-reported benefit (left) or side effects profile (right) for the study medications. Amitriptyline and buprenorphine are excluded due to the lack of variance in their Pillcheck scores (see text).

| medication | prediction correlation with reported benefit |  | prediction correlation with reported harm |  |
| --- | --- | --- | --- | --- |
|  | Spearman's rho | p value | Spearman's rho | p value |
| nortriptyline | -0.08 | 0.96 | 0.06 | 0.83 |
| amitriptyline | * | * | * | * |
| duloxetine | 0.30 | 0.35 | 0.22 | 0.37 |
| tramadol | -0.01 | 0.97 | 0.12 | 0.60 |
| nabilone | -0.54 | 0.67 | -0.62 | 0.29 |
| buprenorphine patch | * | * | * | * |

## Discussion

In this small cohort of pain patients with comorbid psychiatric disorders, genetic profiling using Pillcheck did not seem to correlate with reported benefit or side effect profile of commonly prescribed pain medications. Furthermore, discontinued medications were no more likely to be marked as warranting caution than did actively used medications.

While from the clinicians perspective there could be a benefit for clinical decision making in certain yet better to be defined situations, the questions is whether the cost of testing is worthwhile on a large scale basis [17]. In addition, Leland *et al*. showed that while some clinicians felt well informed about genetic testing, it was mental health providers who were more comfortable ordering pharmacogenetic testing compared to primary providers who mainly did genetic testing for diagnostic purposes [30]. There are also numerous medications commonly used in clinical practice that are not captured by current pharmacogenetic testing which limits conclusion regarding this tool.

Our trial, while unique with respect to the population studied, showed similar results as previous studies. Given the lack of utility for normal metabolizers in the Smith *et al*. trial only a small percentage of the cohort would appear to have benefited from the guided recommendations [25].

Olson *et al*. showed the percentage of patients experiencing side effects was lower using guided therapy, but did recognize that all adverse outcomes cannot be prevented because factors such as age, comorbidities, or other factors can affect drug pharmacokinetics and pharmacodynamics [29]. The nature of the proprietary algorithm used in the Greden trial may impact the generalizability of any of the noted clinical benefits, given the testing used in this trial [26].

The size, lack of control, and retrospective nature of the patient reported data are important limitations of this study. The results also represent medication use at any point, therefore perhaps under the guidance of a specialist, medication selection and dose optimization could have resulted in different perception of side effects and clinical efficacy.

While the lack of observed correlation may also result from an underpowered cohort, other explanations should be considered. While the variability of individual response to medications, including their efficacy and side effects profile, depends upon genetic variability, other factors have been shown to play an important role as well, including drug-drug interactions and the composition and function of the gut microbiome to name just a few [15,16].

Some authors argue that focusing solely on wide scale testing and patient outcomes may not be the best measure of utility of pharmacogenetic testing, as overall healthcare utilization costs may improve with application of testing high risk cases, although the mechanism of benefit would be worthwhile exploring further [28]. Therefore a larger, prospective, controlled study, measuring numerous clinical and non-clinical outcomes would be worthwhile in the future before widespread adoption for patients living with chronic pain. Future studies can also attempt to address the relative importance of factors such as diet, environment, microbiome and disease on the pharmacologic medication processing.

## Conclusion

In this small cohort of pain patients with comorbid psychiatric disorders, genetic profiling using Pillcheck did not seem to correlate with reported benefit or side effect profile of commonly prescribed pain medications. Furthermore, discontinued medications were no more likely to be marked as warranting caution than did actively used medications.

## Data Availability

We have all data available

https://www.healthaya.com/retrospective-analysis

